# Cross-Sectional Analysis of the Prevalence and Risk Factors Associated with Malnutrition Risk Among Oncology Patients Using Two Validated Malnutrition Screening Tools

**DOI:** 10.64898/2026.01.17.26344322

**Authors:** Rachel Hoobler, Manuela Herrera, Kary Woodruff, Adriana M Coletta, Amandine Chaix, Alejandro Sanchez, Joan Elizondo, Thomas K Varghese, Mary C Playdon

## Abstract

**Background:** The prevalence of malnutrition risk among oncology patients across a wide range of cancer types in the United States (US) has rarely been investigated. The objective of this study was to characterize malnutrition risk by demographic and cancer characteristics, and to identify potential risk factors for malnutrition risk in diverse cancer patient populations.

**Methods:** A cross-sectional analysis was conducted using Electronic Medical Record data from 2,894 adult patients living with cancer who underwent malnutrition screening at nine outpatient cancer clinics at the Huntsman Cancer Institute, with a catchment area that includes Utah, Idaho, Montana, Nevada, and Wyoming. Data were collected from January 2021 to April 2024. Malnutrition risk was evaluated using two validated malnutrition screening tools: the Malnutrition Screening Tool (MST) and NUTRISCORE. T-tests and Chi square tests of independence were used to assess differences in sociodemographic, geographic, and cancer characteristics between those with and without malnutrition risk. Logistic regression was implemented to identify potential risk factors for malnutrition risk in an outpatient cancer patient population.

**Results:** Malnutrition risk prevalence for all cancers combined was 23% according to MST and 12% according to NUTRISCORE. Advanced cancer stage and current or previous smoking status were significant risk factors for malnutrition risk regardless of malnutrition screening tool. Head and neck, gastrointestinal, lung cancers, and malignant cancers with an unclear primary site had the highest prevalence of malnutrition risk.

**Conclusions:** Oncology outpatient clinics in the US Mountain West have a malnutrition risk prevalence of up to 23%. Patients with advanced cancer stages, smokers or former smokers, and those diagnosed with head and neck, gastrointestinal, and lung cancers may be at increased risk for malnutrition. Implementation of validated malnutrition screening tools is warranted across outpatient oncology institutions.

## Background

Cancer-related malnutrition, defined by insufficient caloric intake and/or nutrient malabsorption leading to involuntary weight and muscle loss,^1^ poses a serious risk to patients with cancer, as it is associated with worse clinical and survival outcomes.^2–8^ Among oncology patients, malnutrition may be driven by multiple factors including systemic effects of the tumor and side effects of anti-cancer treatments that can negatively affect physical activity and instigate nutrition impact symptoms (e.g., altered taste, reduced appetite, nausea and vomiting, diarrhea, mucositis, constipation, and fatigue).^9^ Ultimately, the combination of these factors increase the risk of weight loss, muscle atrophy, and impaired immunity that can complicate cancer treatment and hinder the recovery process.^10^

Malnutrition status is determined through a multistep process. Patients first undergo malnutrition screening to determine if they are at risk for malnutrition.^11^ Patients determined to be at risk are referred for malnutrition assessment to make an official malnutrition diagnosis followed by medical nutrition therapy as appropriate.^12^ Multiple validated malnutrition screening and malnutrition assessment tools exist for these purposes.^11, 13, 14^

Despite literature demonstrating the importance of malnutrition in patient prognosis, malnutrition screening is not standard of care in about half of outpatient clinics among cancer centers in the US.^15^ Few studies have investigated the prevalence of malnutrition or malnutrition risk across diverse cancer types in the ambulatory setting, and to our knowledge, only one of these studies was conducted in the US.^16^ Implementation of validated malnutrition screening tools is vital for early identification of malnutrition so patients can receive timely nutrition intervention, which can improve the nutritional status of oncology patients.^17^ Enhanced understanding of the prevalence of malnutrition among diverse cancer types as well as potential risk factors for malnutrition is essential for promoting and directing malnutrition screening efforts.

This descriptive study aims to summarize the prevalence of malnutrition risk among patients with any cancer type using malnutrition screening embedded within clinic workflow at nine Huntsman Cancer Institute (**HCI**) outpatient clinics, summarize the sociodemographic, clinical, and geographic characteristics of oncology patients who screened at risk for malnutrition compared to those who were not at risk, and investigate potential risk factors of malnutrition risk.

## Methods

A retrospective cohort study, the Malnutrition Among Cancer Survivors (**MACS**) cohort, was conducted using Electronic Medical Record (**EMR**) data collected from January 2021 to April 2024. This study involved nine outpatient clinics (head and neck, thoracic, gastrointestinal, sarcoma, neurology, bone marrow transplant/hematology, genitourinary, melanoma, and breast/gynecologic) at HCI serving the Mountain West region, including Utah, Idaho, Montana, Nevada, and Wyoming. Included patients were adults 18 years or older with a Malnutrition Screening Tool (**MST**) score within one year of a cancer diagnosis. We excluded for patients with implausibly high body mass index (**BMI**) (>70 kg/m^2^) (**Figure 1**).

**Figure 1.**
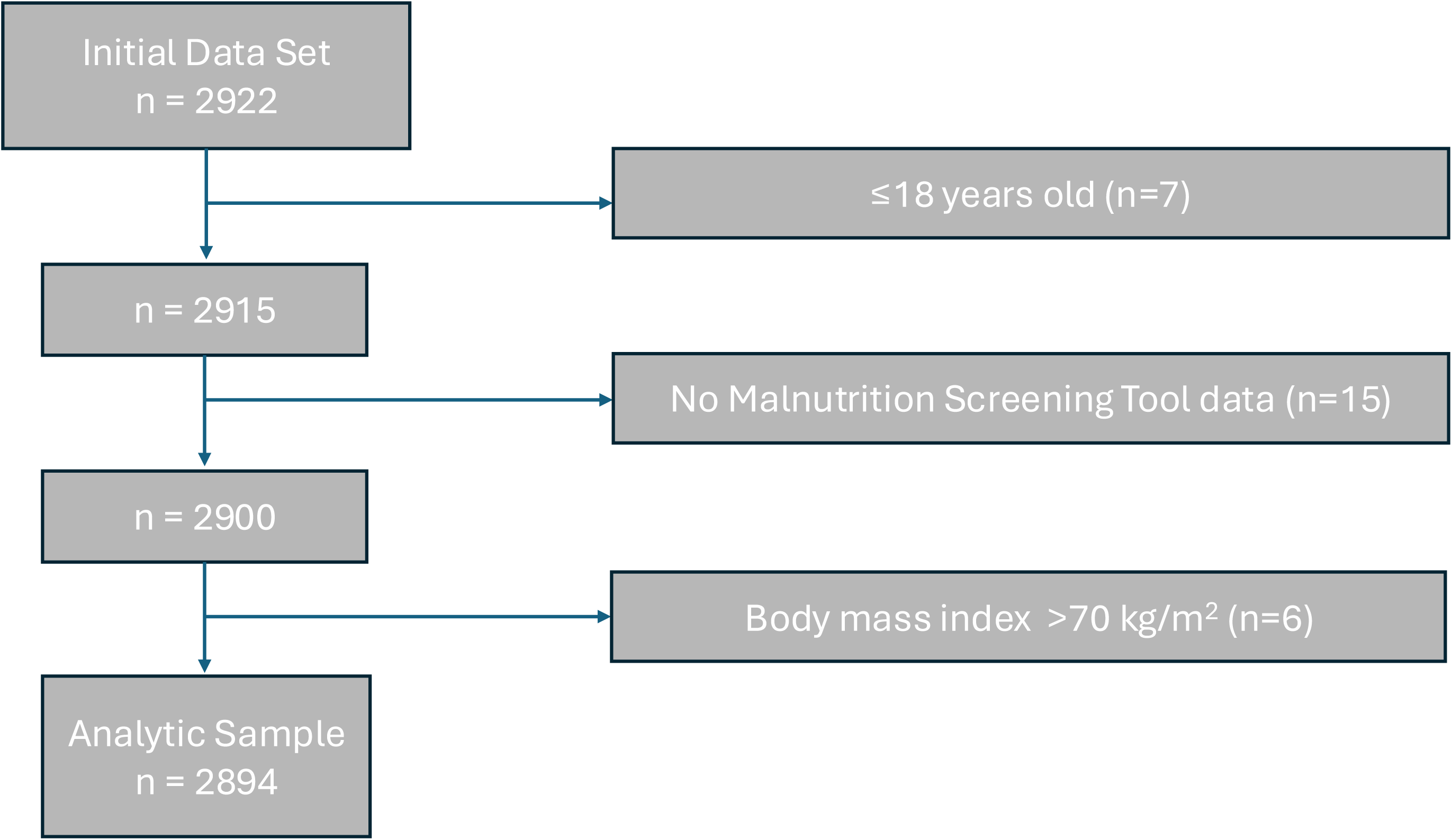
Flow chart of study exclusions.

### Malnutrition Risk

Malnutrition risk was evaluated using MST score available in the EMR and abstracted by the Research Informatics Shared Resource (**RISR**) at HCI.^18^ MST was integrated into standard of care at HCI outpatient clinics in early 2021. Medical assistants screen patients using EPIC Systems EMR prompts every 30 days, unless they have seen, or will see, an outpatient dietitian within 15 days. MST assesses malnutrition risk using patient reported information on changes in appetite or food intake and the presence and extent of unintentional weight loss (**Figure 2**).^18^ A score ≥2 out of 5 indicates malnutrition risk, and a score <2 indicates no malnutrition risk.^14, 18^ MST is a validated malnutrition screening tool for inpatient and outpatient use,^11^ and is estimated to have a sensitivity of 84.0% and a specificity of 85.6% when compared to comprehensive Patient-Generated Subjective Global Assessment (**pg-SGA**).^19^ If patients had more than one MST score available in the EMR, the highest MST score was utilized for this analysis.

**Figure 2.**
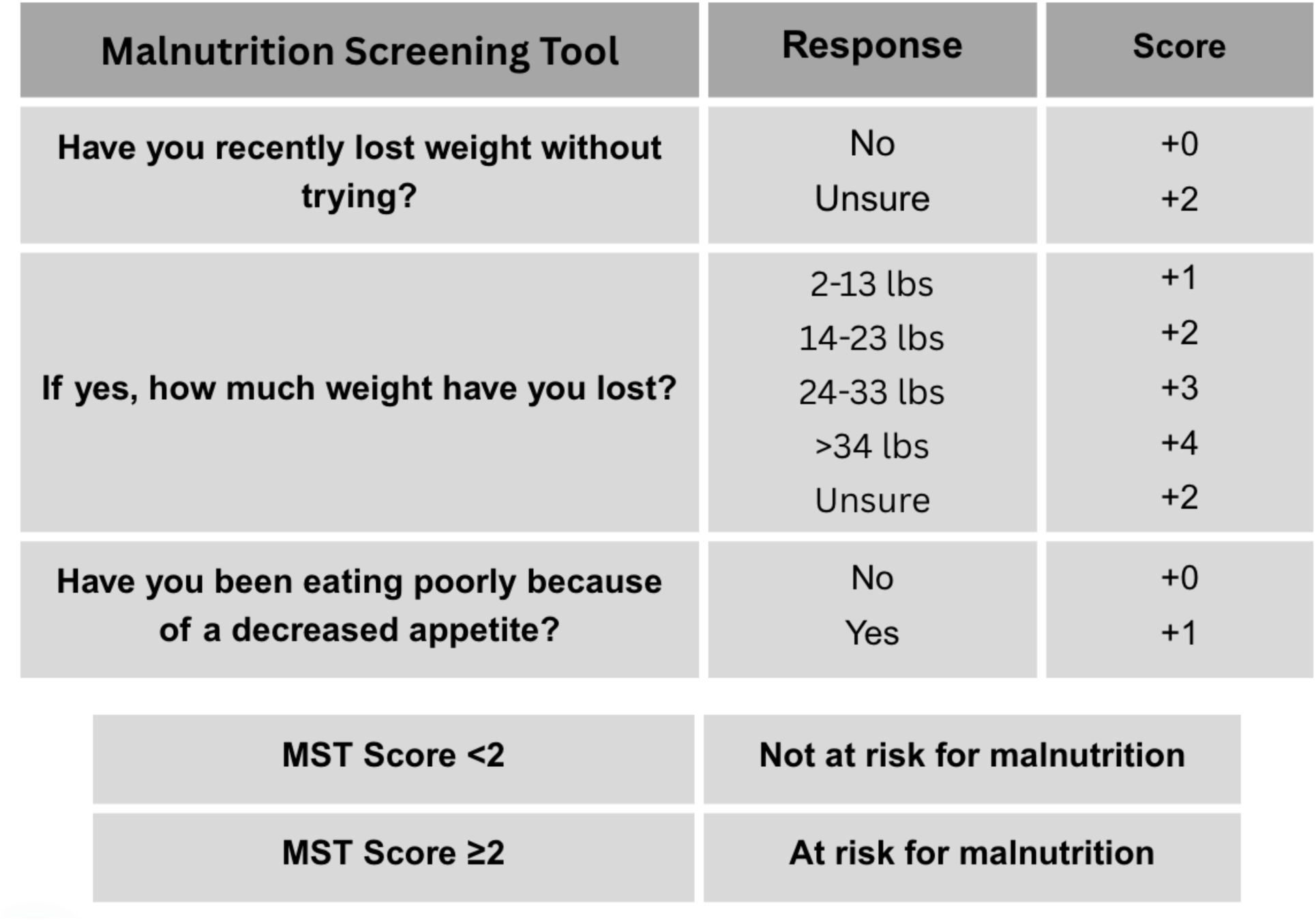
Malnutrition Screening Tool (MST) scoring criteria.

We also assessed malnutrition risk using NUTRISCORE. NUTRISCORE computes malnutrition risk using the same criteria as MST, in addition to considering cancer type and cancer treatment (**Figure 3**).^19^ We included NUTRISCORE as a comparison malnutrition screening tool due to its high estimated sensitivity (97.3%) and specificity (95.9%) when compared to pg-SGA, which exceeds that of MST.^19^ NUTRISCORE scores were calculated for each participant based on the weight loss and appetite data collected as part of MST screening and using the cancer type and cancer treatment data available in the EMR. We were unable to abstract data on hyperfractionated radiation therapy and hematologic stem cell transplantation from the EMR; therefore, these treatments were excluded from our scoring criteria. A NUTRISCORE score ≥5 indicates malnutrition risk and a score <5 indicates no risk.^14, 19^ We again selected each patient’s highest NUTRISCORE score when multiple scores were available.

**Figure 3.**
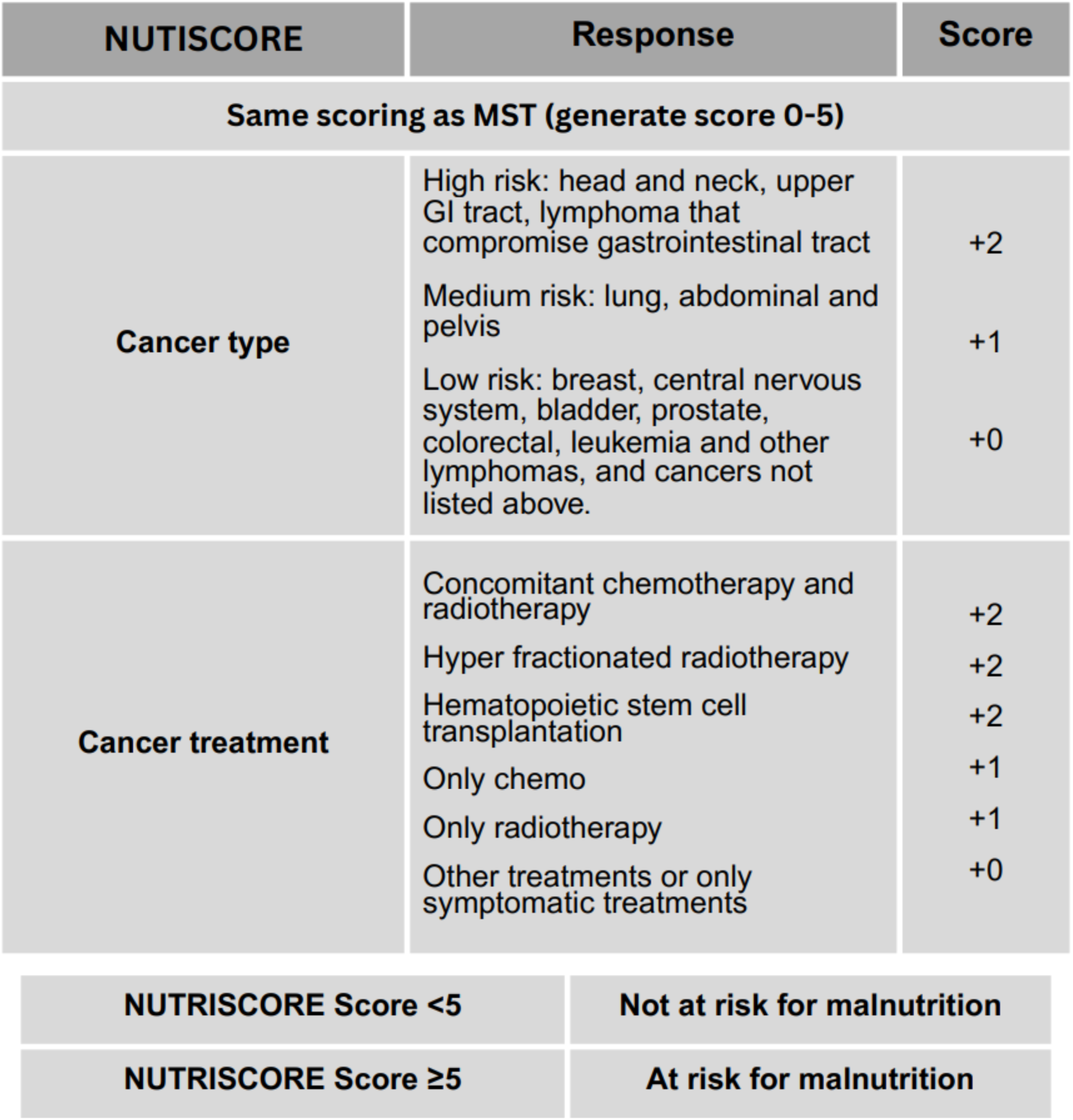
NUTRISCORE scoring criteria.

### Sociodemographic and Cancer Characteristics

Sociodemographic and cancer characteristics were obtained from medical records, including age at cancer diagnosis (continuous), biologic sex (female, male), race (white, non-white (i.e., Black or African American, American Indian or Alaska Native, Asian, Native Hawaiian or Pacific Islander), missing), ethnicity (non-Hispanic, Hispanic/Latino, missing), marital status (married, unmarried, missing), smoking status (never, quit, current), rural/urban status (rural, urban), BMI at first MST score, cancer stage (0/I, II, III, IV), and cancer treatment (surgery, chemotherapy, radiation, immunotherapy, and hormone therapy). Age over 90 years was imputed as 90 years due to Health Insurance Portability and Accountability Act (**HIPAA**) protections.^20^ Zip code was used to determine rural or urban residence using Rural-Urban Commuting Area (**RUCA**) codes, with codes 1-6 categorized as urban and 7-10 as rural.^21^ BMI was categorized as underweight (BMI <18.5 kg/m^2^), normal weight (BMI 18.5-24.99 kg/m^2^), overweight (BMI 25-29.99 kg/m^2^), or obese (BMI >30 kg/m^2^), using World Health Organization (**WHO**) criteria.^22^

Cancer type was ascertained using International Classification of Diseases, for Oncology (**ICD-O**) codes (**Supplementary Table 1**). The ICD-O classification codes for head and neck had cross-classification with non-melanoma skin cancers of the lip, ear, scalp and neck, and other unspecified parts of the face (C44.0, C44.2, C44.3, C44.4). For the purposes of this analysis, these ICD-O codes were categorized as head and neck cancers. We omitted cancers related to metastatic spread, such as secondary lymph node cancer (ICD-O, C77) and malignant cancers with unknown or ill-defined primary site (ICD-O, C76 and C80), unless these were a patient’s only cancer diagnosis in the EMR within one year of an MST score. In the case of patients with multiple cancers, we prioritized the cancer type with the highest stage, as cancer stage has consistently been associated with increased risk for malnutrition.^23, 24^ If two or more cancers were of the same stage, we selected the cancer diagnosis closest to the patient’s highest MST score.

### Statistical Analysis

Sociodemographic and cancer characteristics were summarized using median and interquartile range for continuous variables and frequency and percentage per category for categorical variables. We stratified characteristics by malnutrition risk, or no risk, based on MST (MST score ≥2 versus <2) and NUTRISCORE (NUTRISCORE score ≥5 versus <5) criteria.^14^ We compared differences in sociodemographic, cancer, and geographic characteristics using Mann-Whitney U Test for continuous variables, owing to skewed distribution based on Shapiro-Wilk test, and Chi Square Test of Independence for categorical variables.

Univariate logistic regression was used to identify variables individually associated with malnutrition risk. Variables statistically significant in univariate logistic regression (p<0.05) were included in multivariable logistic regression analysis. Variables that reached statistical significance in multivariable analysis were considered potential predictors of malnutrition risk. Chemotherapy and radiation were not considered as potential predictors of malnutrition risk assessed using NUTRISCORE since these are components of the scoring criteria. As several cancer types had a limited number of patients, we did not include cancer type in our regression models to avoid issues with model non-convergence. We described malnutrition risk prevalence stratified by cancer type to provide insight into cancer types where malnutrition risk may be more common. All analyses were conducted at a significance level of p<0.05 and were conducted in R statistical software.^25^

## Results

Demographic and cancer characteristics of the MACS cohort (n = 2,894) are presented in **Table 1**. On average, patients had a median age of 65 years old, with slightly more than half being male (52.0%). Most patients were white (81.9%), married (64.9%), never smokers (60.0%), and resided in urban geographic regions (89.7%). Majority of patients had either overweight (30.7%) or obesity (34.4%). Approximately one-third of patients were diagnosed with stage 0/I cancer, followed by stage IV cancer (21.7%), while smaller proportions had stage II (13.5%) or stage III (14.8%) cancer. Over half of patients in the MACS cohort underwent surgery (57.4%), 40.9% received chemotherapy, 28.1% received radiation, and a small portion were treated with either immunotherapy (18.5%) or hormone therapy (14.1%). On average, treatment occurred within a year of patients’ highest MST score.

**Table 1.** Demographic and clinical characteristics of patients with cancer treated at the Huntsman Cancer Institute outpatient clinics who underwent malnutrition screening (n=2,894)

| **Characteristic** | **Median (IQR) or Frequency (%)** |
| --- | --- |
| **Age at Diagnosis** | 65.0 (20.0) |
| **Biologic Sex** |  |
| Female | 1390 (48.0%) |
| Male | 1504 (52.0%) |
| **Race** |  |
| White | 2370 (81.9%) |
| Non-white | 230 (7.9%) |
| **Ethnicity** |  |
| Non-Hispanic | 2475 (85.5%) |
| Hispanic or Latino | 188 (6.5%) |
| **Marital Status** |  |
| Married | 1877 (64.9%) |
| Unmarried | 907 (31.3%) |
| **Smoking Status** |  |
| Never | 1735 (60.0%) |
| Quit | 941 (32.5%) |
| Current | 218 (7.5%) |
| **Urban/Rural Status** |  |
| Urban | 2597 (89.7%) |
| Rural | 297 (10.3%) |
| **Body Mass Index (kg/m<sup>2</sup>)** |  |
| Underweight | 56 (1.9%) |
| Normal Weight | 848 (29.3%) |
| Overweight | 888 (30.7%) |
| Obese | 995 (34.4%) |
| **Cancer Stage** |  |
| 0/I | 870 (30.1%) |
| II | 390 (13.5%) |
| III | 428 (14.8%) |
| IV | 627 (21.7%) |
| Missing |  |
| **Surgery** |  |
| No | 1232 (42.6%) |
| Yes | 1662 (57.4%) |
| **Chemotherapy** |  |
| No | 1709 (59.1%) |
| Yes | 1185 (40.9%) |
| **Radiotherapy** |  |
| No | 2080 (71.9%) |
| Yes | 814 (28.1%) |
| **Immunotherapy** |  |
| No | 2359 (81.5%) |
| Yes | 535 (18.5%) |
| **Hormone Therapy** |  |
| No | 2487 (85.9%) |
| Yes | 407 (14.1%) |
| Percentages may not add up to 100% due to missing data. |  |

### Overall Malnutrition Risk

655 patients (22.6%) were at risk for malnutrition using MST criteria (MST score ≥2). According to NUTRISCORE classification, 337 patients (11.6%) were at risk for malnutrition (NUTRISCORE score ≥5).

### Patient Characteristics Stratified by Malnutrition Risk

**Table 2** presents demographic and cancer characteristics of patients in the MACS cohort stratified by malnutrition risk. Patients at risk for malnutrition measured by MST were more likely to be male, older, current/former smokers, had lower BMI, higher cancer stage, and received chemotherapy, radiation, or immunotherapy. In contrast, a smaller proportion of patients who underwent surgery or hormone therapy were at risk for malnutrition.

**Table 2.**
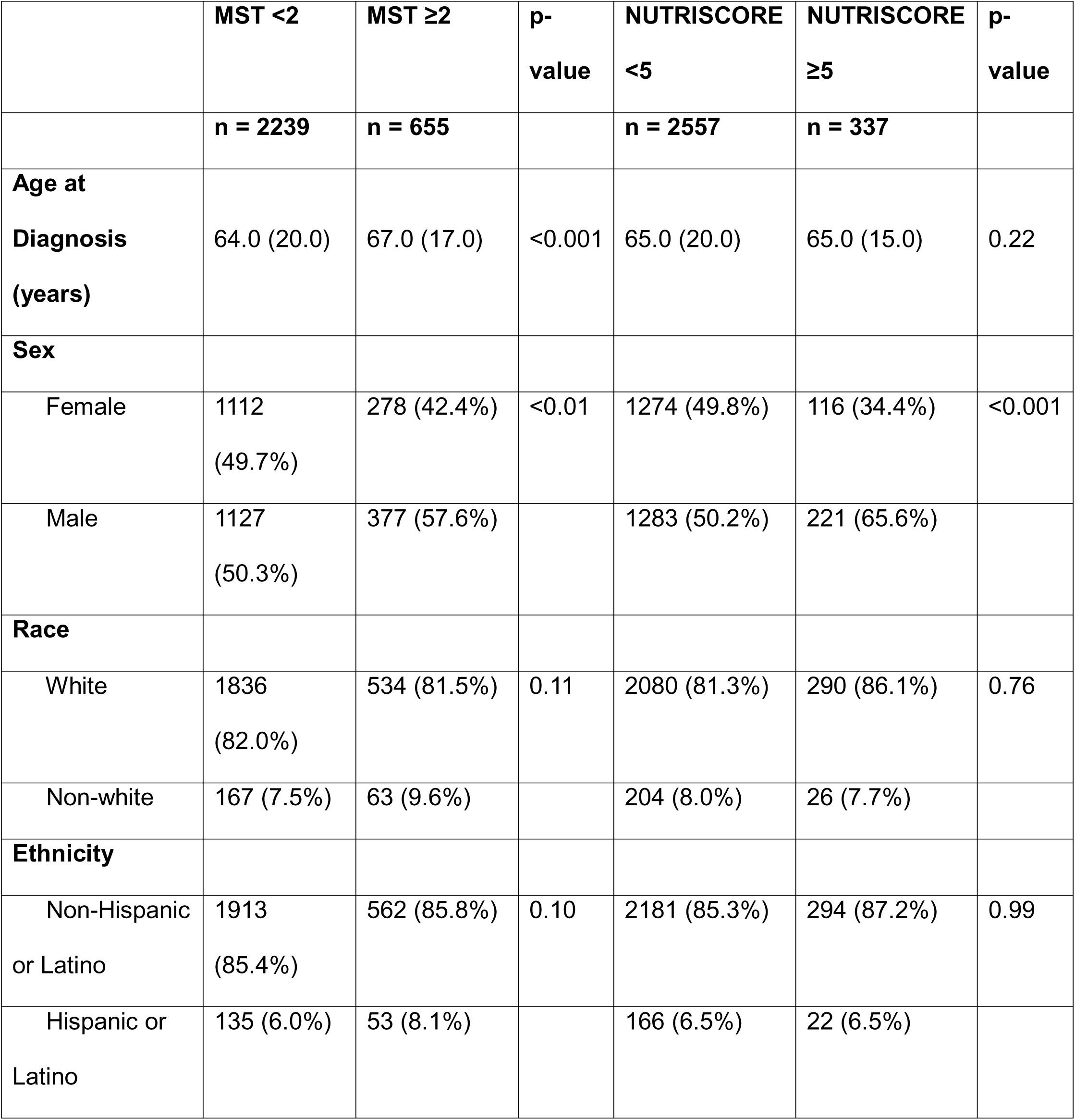
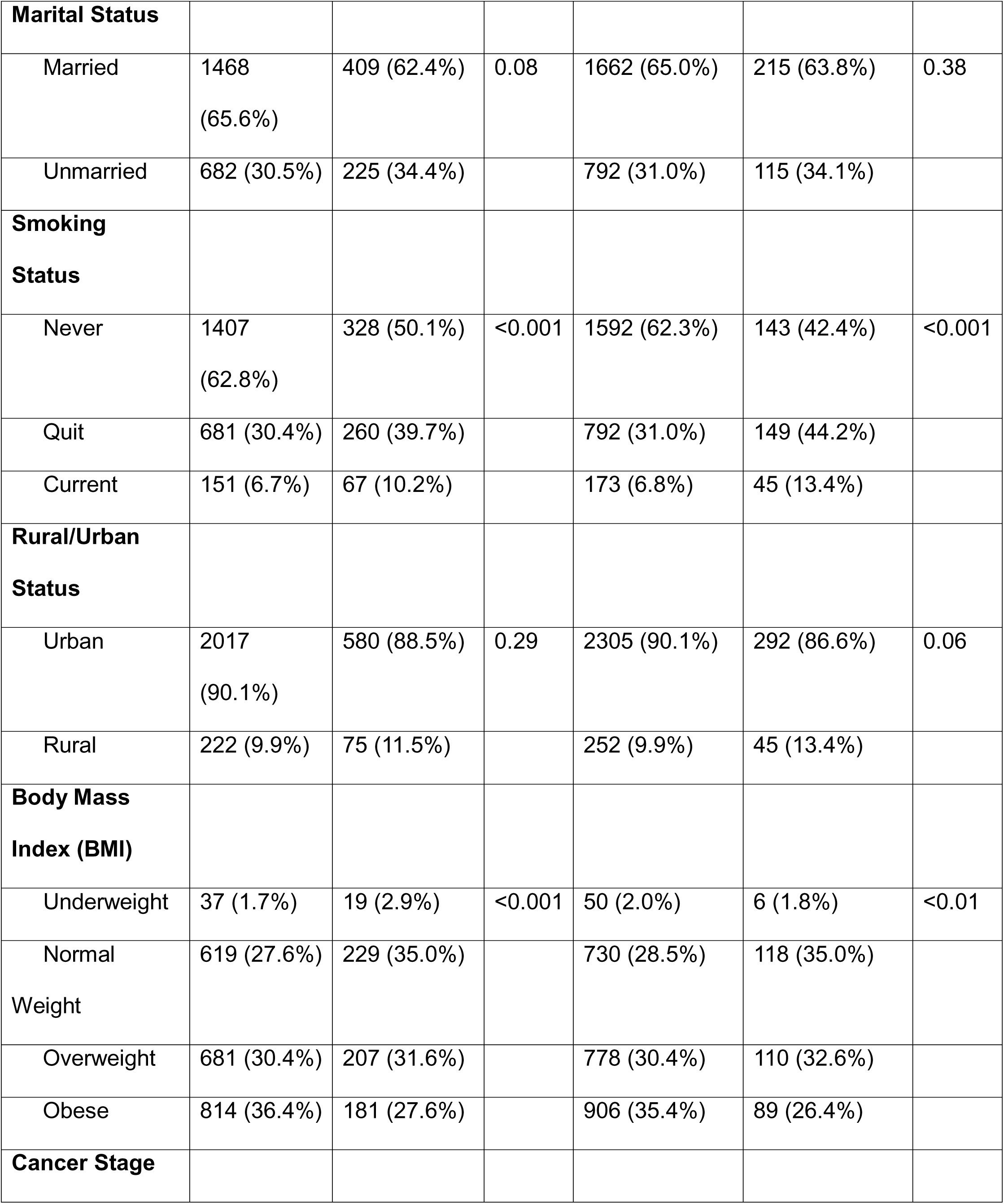
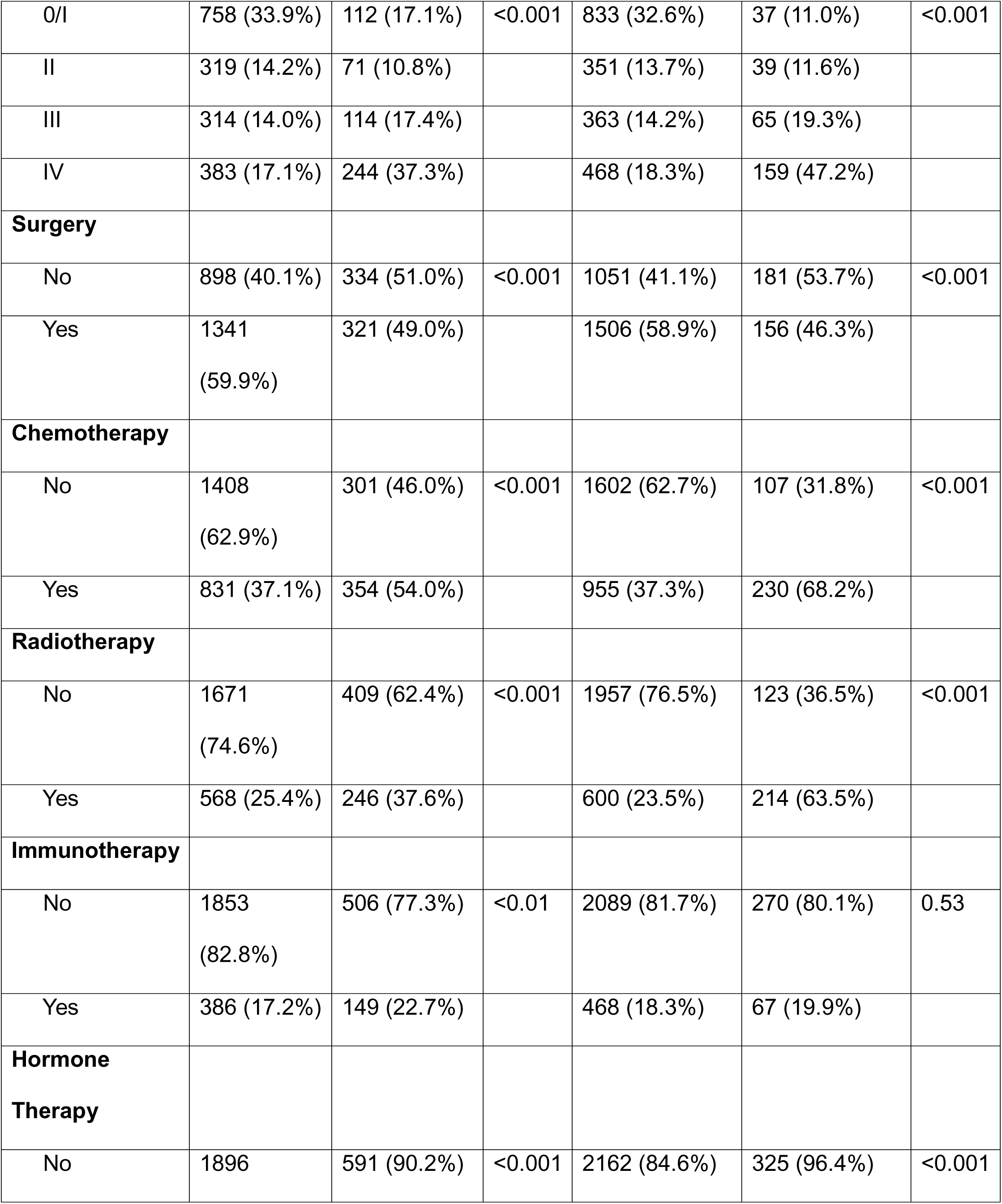
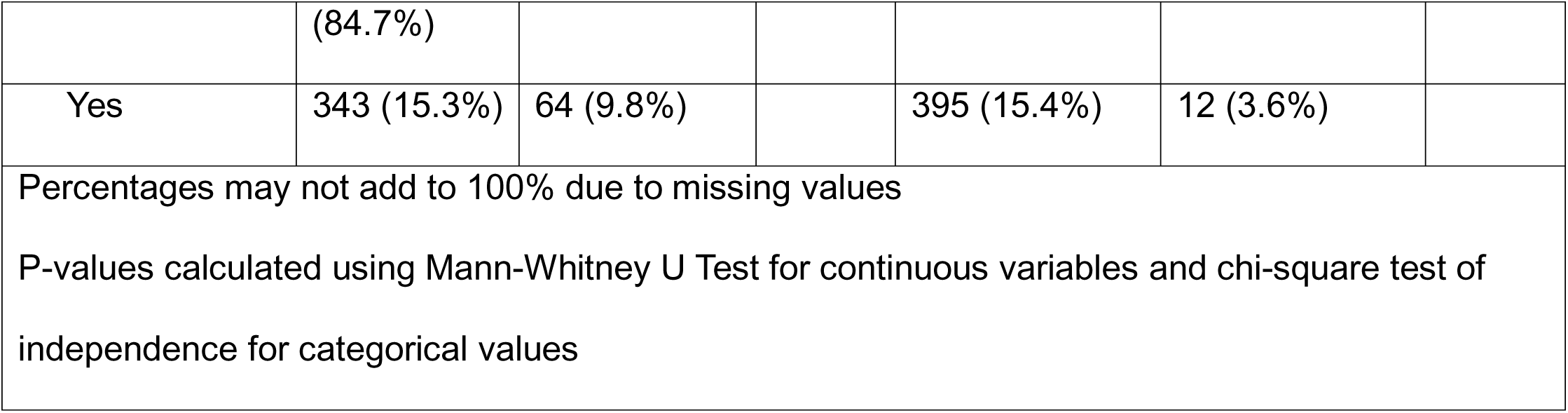
Demographic and clinical characteristics of patients with cancer treated at the Huntsman Cancer Institute outpatient clinics who underwent malnutrition screening (n=2,894) stratified by malnutrition risk determined by Malnutrition Screening Tool (MST) score and NUTRISCORE.

Those at risk for malnutrition according to NUTRISCORE were also more likely to be male, current/former smokers, have advanced disease, and less likely to have obesity. There was a higher proportion of malnutrition risk among patients treated with chemotherapy and radiation, and a lower proportion of malnutrition risk among patients who underwent surgery or received hormone therapy.

### Predictors of Malnutrition Risk

Univariate logistic regression identified that older age, male sex, being a former or current smoker, stage III or IV cancer, and treatment with chemotherapy, radiation or immunotherapy were associated with increased risk for malnutrition risk assessed using MST among oncology patients in the MACS cohort. Obesity and treatment with surgery or hormone therapy were associated with reduced risk for malnutrition risk assessed using MST. In multivariable logistic regression, older age, former/current smoking status, stage III and IV cancer, and treatment with chemotherapy or radiation remained significantly associated with increased risk for malnutrition risk. Obesity remained inversely associated with malnutrition risk defined by MST (**Table 3**).

**Table 3.** Association of demographic and cancer characteristics with malnutrition risk defined by Malnutrition Screening Tool (MST) (MST score ≥2 versus <2) using univariable and multivariable logistic regression among adult cancer patients treated at the Huntsman Cancer Institute outpatient clinics (n=2,894)

| Characteristic | Univariate |  | Multivariable |  |
| --- | --- | --- | --- | --- |
|  | OR (95% CI) | P-Value | OR (95% CI) | P-Value |
| Age | 1.01 (1.01-1.02) | $<0.001$ | 1.01 (1.00-1.02) | $<0.001$ |
| Biologic Sex |  |  |  |  |
| Female | 1.00 (Ref) |  | 1.00 (Ref) |  |
| Male | 1.34 (1.12-1.60) | $<0.001$ | 1.17 (0.97-1.41) | 0.10 |
| Race |  |  |  |  |
| White | 1.00 (Ref) |  |  |  |
| Non-White | 1.30 (0.96-1.76) | 0.10 |  |  |
| Ethnicity |  |  |  |  |
| Non-Hispanic or Latino | 1.00 (Ref) |  |  |  |
| Hispanic or Latino | 1.34 (0.96-1.86) | 0.09 |  |  |
| Marital Status |  |  |  |  |
| Married | 1.00 (Ref) |  |  |  |
| Unmarried | 1.18 (0.98-1.43) | 0.08 |  |  |
| Smoking Status |  |  |  |  |
| No | 1.00 (Ref) |  | 1.00 (Ref) |  |
| Quit | 1.64 (1.36-1.97) | $<0.001$ | 1.40 (1.15-1.71) | $<0.001$ |
| Current | 1.90 (1.39-2.60) | $<0.001$ | 1.57 (1.12-2.19) | $<0.01$ |
| Body Mass Index (BMI) |  |  |  |  |
| Underweight | 1.39 (0.78-2.46) | 0.26 | 1.23 (0.67-2.26) | 0.50 |
| Normal Weight | 1.00 (Ref) |  | 1.00 (Ref) |  |
| Overweight | 0.82 (0.66-1.02) | 0.08 | 0.90 (0.71-1.12) | 0.34 |
| Obese | 0.60 (0.48-0.75) | <0.001 | 0.72 (0.57-0.92) | <0.01 |
| Rural/Urban Status |  |  |  |  |
| Urban | 1.00 (Ref) |  |  |  |
| Rural | 1.17 (0.89-1.55) | 0.26 |  |  |
| Cancer Stage |  |  |  |  |
| 0/I | 1.00 (Ref) |  | 1.00 (Ref) |  |
| II | 1.51 (1.09-2.08) | 0.01 | 1.19 (0.85-1.66) | 0.32 |
| III | 2.46 (1.83-3.29) | <0.001 | 1.76 (1.28-2.41) | <0.001 |
| IV | 4.31 (3.34-5.57) | <0.001 | 3.17 (2.37-4.23) | <0.001 |
| Surgery |  |  |  |  |
| No | 1.00 (Ref) |  | 1.00 (Ref) |  |
| Yes | 0.64 (0.54-0.77) | <0.001 | 1.04 (0.85-1.28) | 0.69 |
| Chemotherapy |  |  |  |  |
| No | 1.00 (Ref) |  | 1.00 (Ref) |  |
| Yes | 1.99 (1.67-2.38) | <0.001 | 1.55 (1.25-1.92) | <0.001 |
| Radiation |  |  |  |  |
| No | 1.00 (Ref) |  | 1.00 (Ref) |  |
| Yes | 1.77 (1.47-2.13) | <0.001 | 1.56 (1.28-1.90) | <0.001 |
| Immunotherapy |  |  |  |  |
| No | 1.00 (Ref) |  | 1.00 (Ref) |  |
| Yes | 1.41 (1.14-1.75) | <0.001 | 0.85 (0.67-1.09) | 0.21 |
| Hormone Therapy |  |  |  |  |
| No | 1.00 (Ref) |  | 1.00 (Ref) |  |
| Yes | 0.60 (0.45-0.79) | <0.001 | 0.79 (0.58-1.07) | 0.13 |

Male sex, former/current smoking status, rural residence, and stage II, III, and IV cancer were univariately positively associated with risk for malnutrition defined by NUTRISCORE. Obesity, surgery, and hormone therapy were inversely associated with malnutrition risk. Obesity, rural residence, and surgery were not retained in the multivariable logistic regression (**Table 4**).

**Table 4.** Association of demographic and cancer characteristics with malnutrition risk defined by NUTRISCORE (NUTRISCORE score ≥5 versus <5) using univariable and multivariable logistic regression among adult cancer patients treated at the Huntsman Cancer Institute outpatient clinics (n=2,894)

| Characteristic | Univariate |  | Multivariable |  |
| --- | --- | --- | --- | --- |
|  | OR (95% CI) | P-Value | OR (95% CI) | P-Value |
| Age | 1.01 (1.00-1.02) | 0.08 |  |  |
| Biologic Sex |  |  |  |  |
| Female | 1.00 (Ref) |  | 1.00 (Ref) |  |
| Male | 1.89 (1.49-2.40) | <0.001 | 1.57 (1.22-2.02) | <0.001 |
| Race |  |  |  |  |
| White | 1.00 (Ref) |  |  |  |
| Other | 0.91 (0.60-1.40) | 0.68 |  |  |
| Ethnicity |  |  |  |  |
| Non-Hispanic or Latino | 1.00 (Ref) |  |  |  |
| Hispanic or Latino | 0.98 (0.62-1.56) | 0.94 |  |  |
| Marital Status |  |  |  |  |
| Married | 1.00 (Ref) |  |  |  |
| Unmarried | 1.12 (0.88-1.43) | 0.35 |  |  |
| Smoking Status |  |  |  |  |
| No | 1.00 (Ref) |  | 1.00 (Ref) |  |
| Quit | 2.09 (1.64-2.68) | <0.001 | 1.67 (1.29-2.17) | <0.001 |
| Current | 2.90 (2.00-4.19) | <0.001 | 2.23 (1.50-3.32) | <0.001 |
| Body Mass Index (BMI) |  |  |  |  |
| Underweight | 0.74 (0.31-1.77) | 0.50 | 0.49 (0.20-1.23) | 0.13 |
| Normal Weight | 1.00 (Ref) |  | 1.00 (Ref) |  |
| Overweight | 0.87 (0.66-1.16) | 0.35 | 1.03 (0.77-1.39) | 0.83 |
| Obese | 0.61 (0.45-0.81) | <0.001 | 0.81 (0.60-1.11) | 0.19 |
| Rural/Urban Status |  |  |  |  |
| Urban | 1.00 (Ref) |  | 1.00 (Ref) |  |
| Rural | 1.41 (1.00-1.98) | 0.05 | 1.31 (0.91-1.88) | 0.15 |
| Cancer Stage |  |  |  |  |
| 0/I | 1.00 (Ref) |  | 1.00 (Ref) |  |
| II | 2.50 (1.57-3.99) | <0.001 | 2.22 (1.38-3.56) | <0.001 |
| III | 4.03 (2.64-6.15) | <0.001 | 3.22 (2.09-4.97) | <0.001 |
| IV | 7.65 (5.26-11.13) | <0.001 | 6.38 (4.29-9.49) | <0.001 |
| Surgery |  |  |  |  |
| No | 1.00 (Ref) |  | 1.00 (Ref) |  |
| Yes | 0.60 (0.48-0.76) | <0.001 | 0.87 (0.68-1.13) | 0.30 |
| Immunotherapy |  |  |  |  |
| No | 1.00 (Ref) |  |  |  |
| Yes | 1.11 (0.83-1.47) | 0.48 |  |  |
| Hormone Therapy |  |  |  |  |
| No | 1.00 (Ref) |  | 1.00 (Ref) |  |
| Yes | 0.20 (0.11-0.36) | <0.001 | 0.26 (0.14-0.48) | <0.001 |

### Malnutrition Risk Prevalence by Cancer Type

Cancer types with the highest number of patients at risk for malnutrition defined by MST were head and neck (n=165), lung (n=120), gastrointestinal (n=106), and colorectal (n=66) cancers (**Figure 4**). As a proportion of the total number of patients diagnosed with specific cancer types, the highest prevalence of malnutrition risk was observed for malignant cancers with unknown or ill-defined primary site (43.5%), gastrointestinal (37.3%), head and neck (32.7%), and lung (27.1%) cancers (**Figure 5**). Breast (11.6%), non-melanoma skin (10.6%), and male genital (6.5%) cancers had the lowest proportions of patients with malnutrition risk. Similar rankings were observed when malnutrition risk was defined using NUTRISCORE (**Figures 6 and 7**), except no patients with heart, eye and orbit, or bone and joint cancer types were at risk for malnutrition using the NUTRISCORE scoring criteria. Complete results are available in **Supplementary Tables 2-5**.

**Figure 4.**
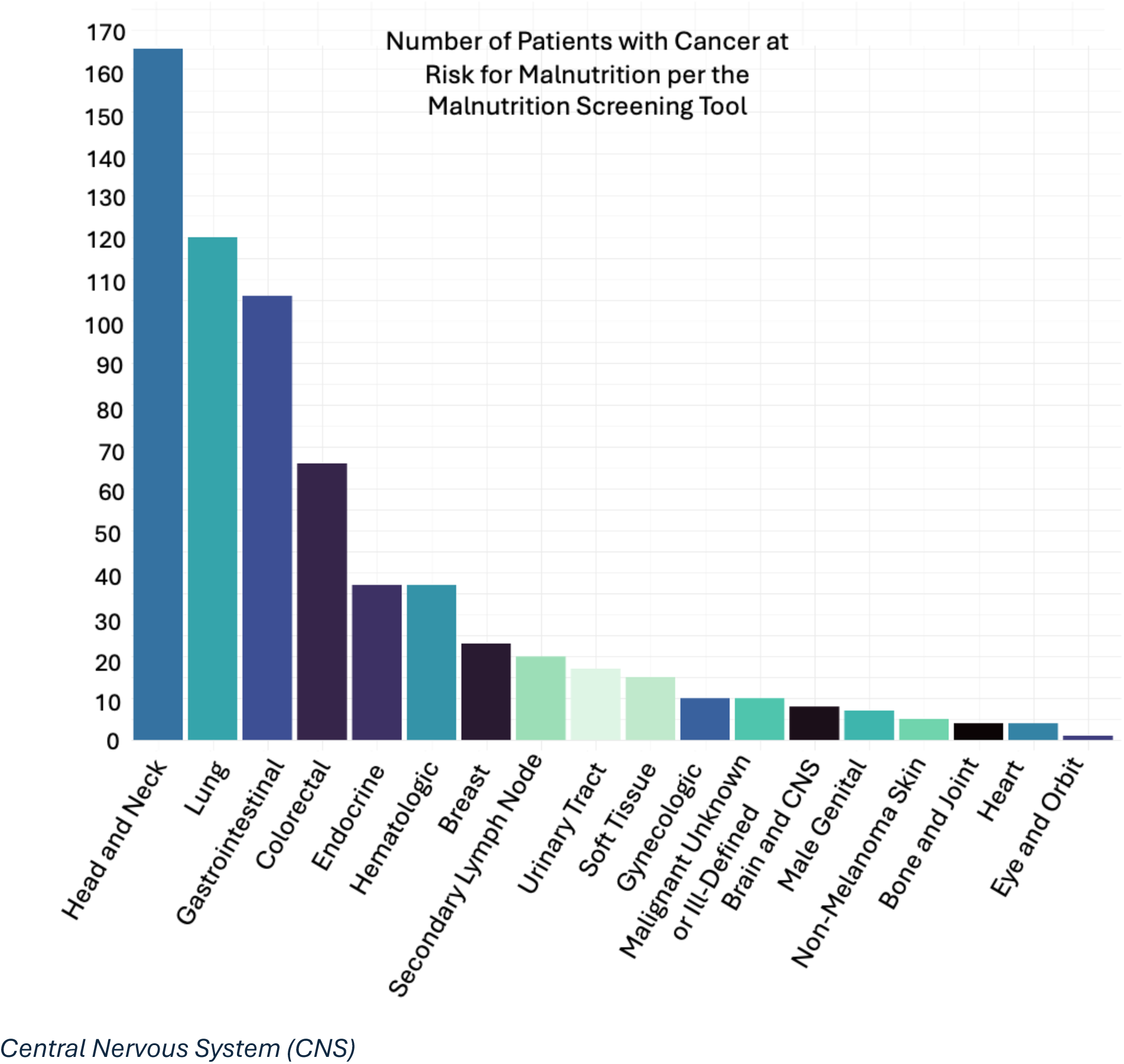
Number of patients with cancer treated at the Huntsman Cancer Institute outpatient clinics at risk for malnutrition using the Malnutrition Screening Tool (MST) by cancer type

**Figure 5.**
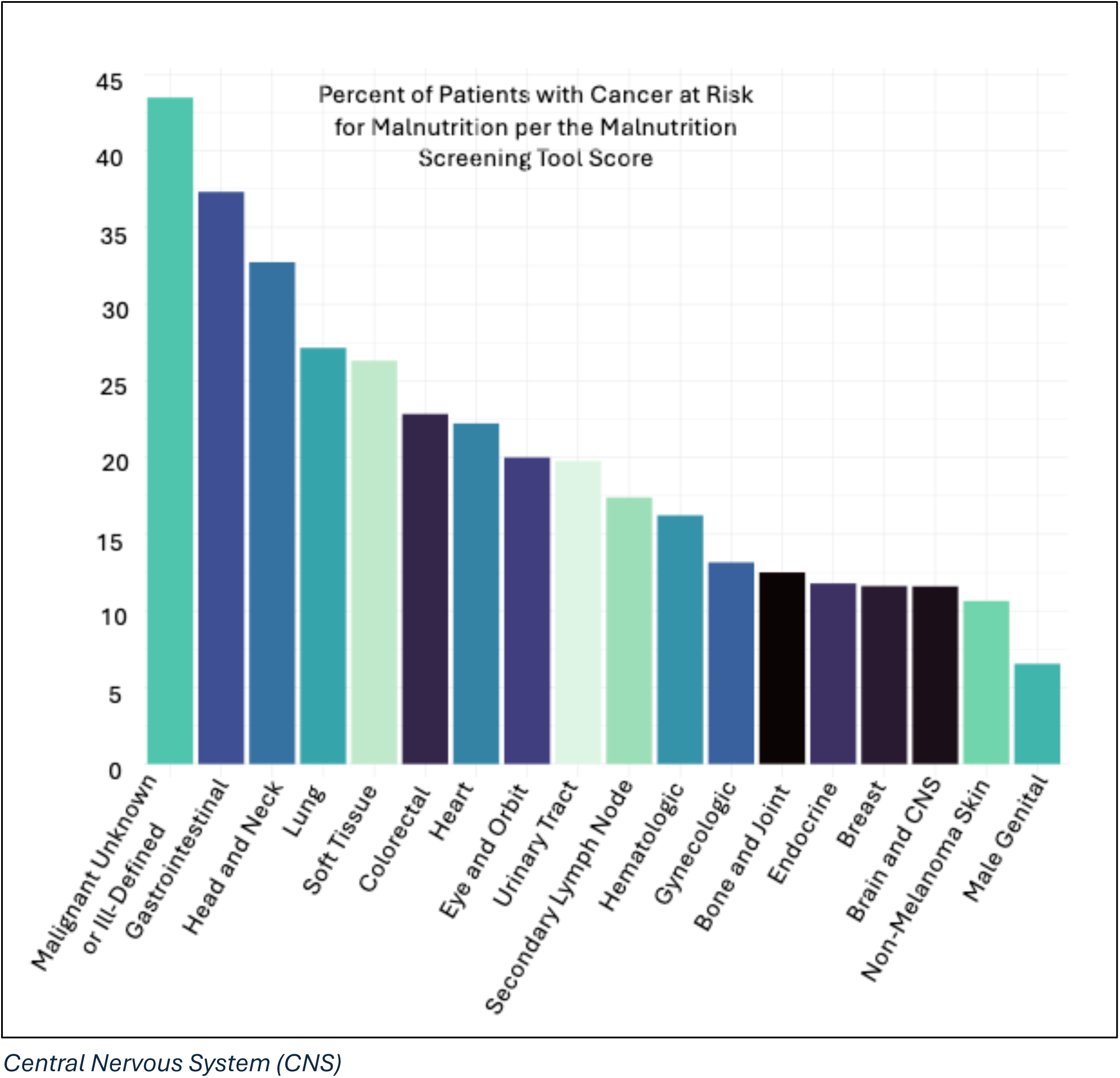
Percent of patients with cancer treated at the Huntsman Cancer Institute outpatient clinics at risk for malnutrition using the Malnutrition Screening Tool (MST) by cancer type

**Figure 6.**
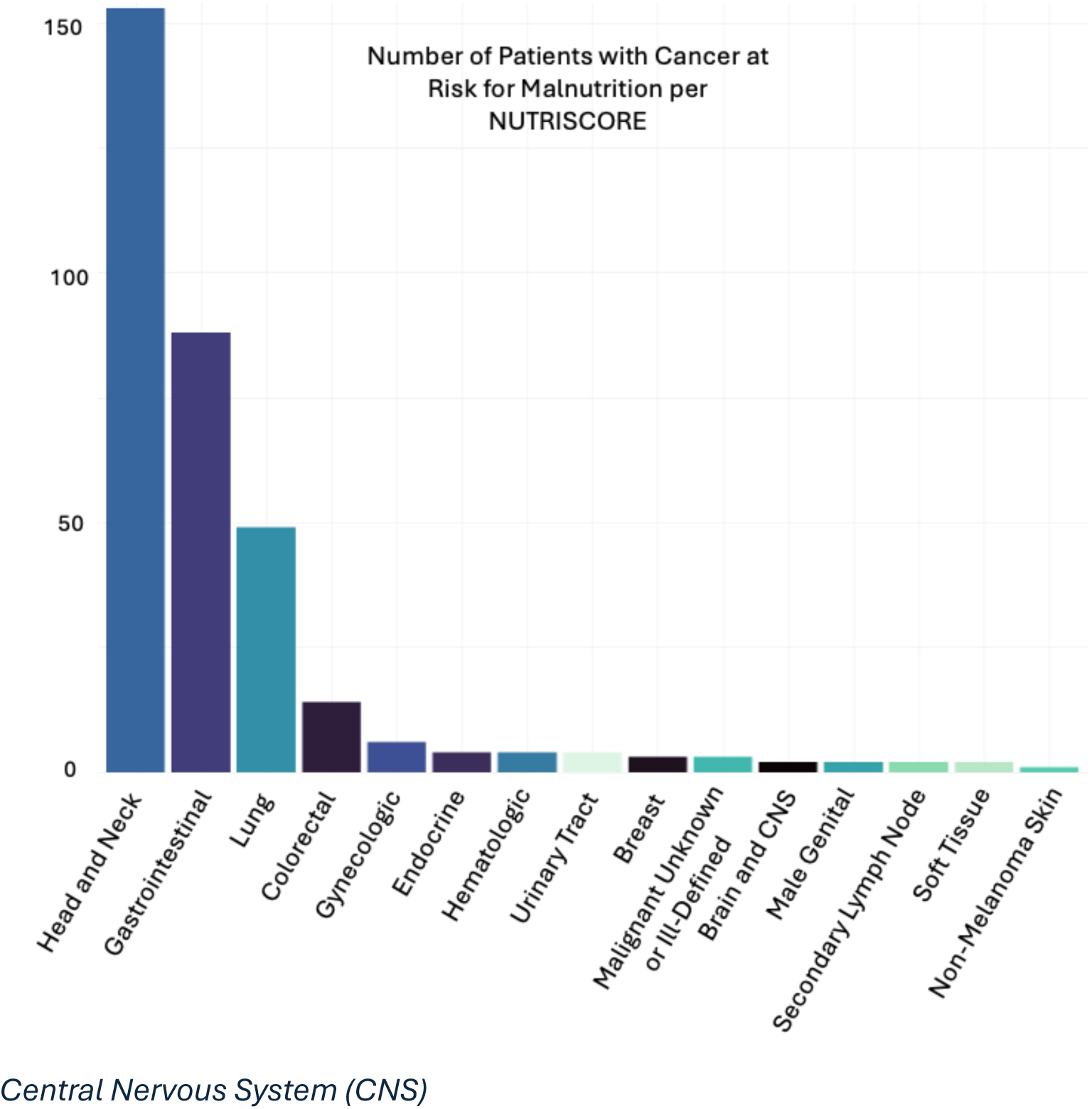
Number of patients with cancer treated at the Huntsman Cancer Institute outpatient clinics at risk for malnutrition using NUTRISCORE by cancer type

**Figure 7.**
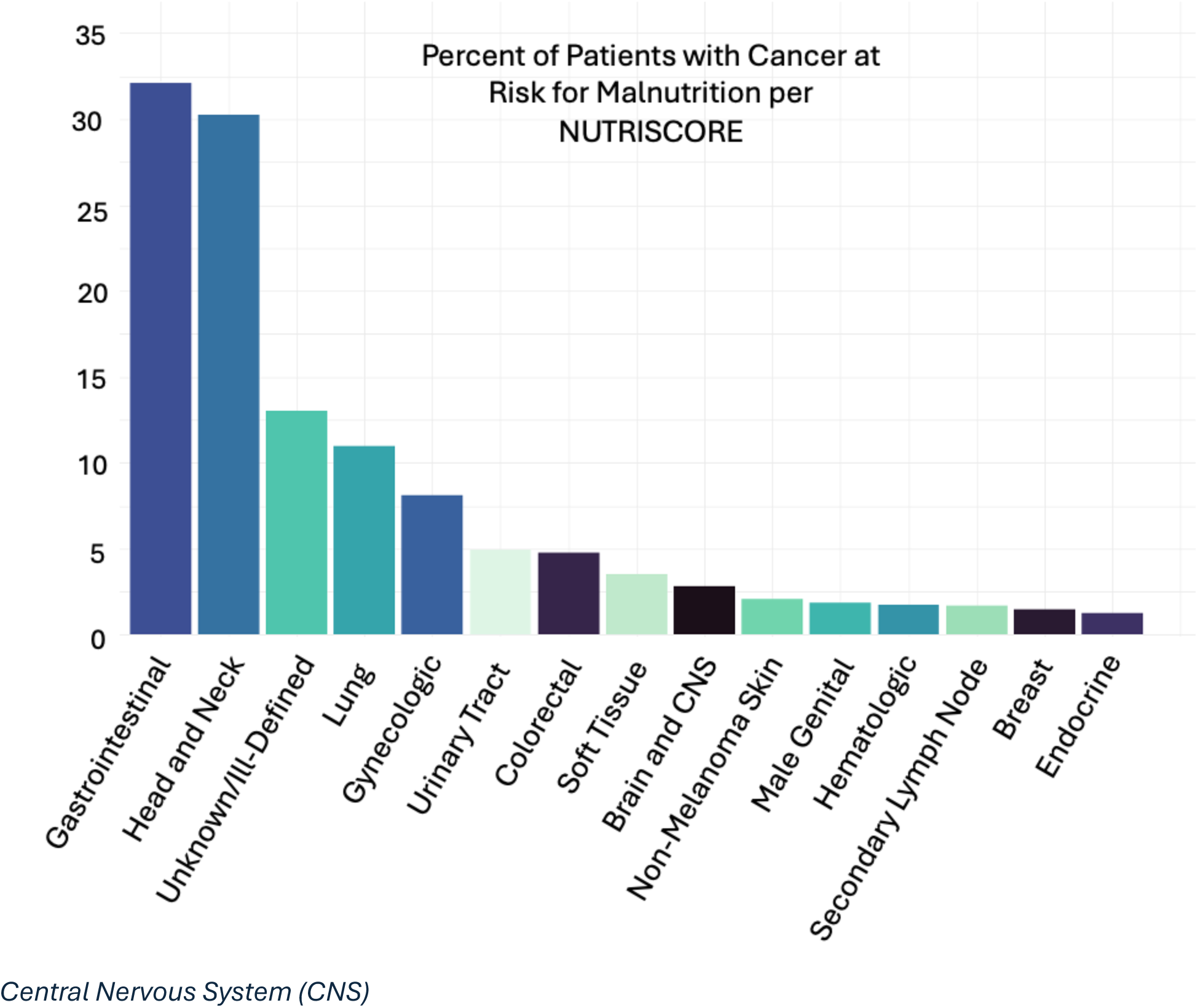
Percent of patients with cancer treated at the Huntsman Cancer Institute outpatient clinics at risk for malnutrition using NUTRISCORE by cancer type

## Discussion

We analyzed the sociodemographic and cancer characteristics of oncology patients who underwent MST screening at HCI, investigated potential risk factors for malnutrition risk, and assessed the period prevalence of malnutrition risk by cancer type over three years using two validated malnutrition screening tools. Twenty-three percent of patients were at risk for malnutrition according to MST, compared to 12% identified by NUTRISCORE. Advanced cancer stage and former or current smoking status were potential risk factors for malnutrition risk measured by both tools. The cancer types with the highest prevalence of malnutrition risk were gastrointestinal, head and neck, and lung cancers, as well as malignant cancers with an unknown or ill-defined primary site.

Previous studies conducted outside of the US have observed malnutrition risk prevalence ranging from 20-30% in the ambulatory oncology setting across multiple cancer types.^26–29^ Only one of these studies used NUTRISCORE to assess malnutrition risk^29^ and none used MST. A recent study in the US found that 28% of outpatient oncology patients were at risk for malnutrition measured using MST among patients with solid tumors in clinics across North and South Carolina, which aligns with our estimates.^16^ Additional studies in the US using validated malnutrition assessment tools, rather than screening tools, are needed to better characterize the prevalence of malnutrition at oncology outpatient clinics.

Current or former smoking status and advanced cancer stage were independent risk factors for malnutrition risk. Our observations are consistent with prior studies that emphasize a high prevalence of malnutrition and implementation of nutrition support among patients with a smoking history.^30–32^ Nicotine has appetite-suppressing effects, which may enhance anorexia.^33^ Moreover, smoking has been linked to lower body weight,^34, 35^ low muscle mass,^36^ and in the US is more common among those of lower income and education level,^37^ all of which could predispose patients to malnutrition. Advanced cancer stage has also been consistently associated with malnutrition and malnutrition risk in previous investigations conducted among oncology patients receiving anti-cancer treatments.^16, 27, 32, 38^ This is likely related to higher rates of cachexia and metabolic derangements at more advanced cancer stages, which drive malnutrition through reduced appetite, increased energy expenditure, and upregulation of pathways that promote muscle atrophy.^39^

Older age at cancer diagnosis was a risk factor for malnutrition risk when assessed by MST but not NUTRISCORE. Older age has regularly been identified as a predictor of malnutrition and malnutrition risk among oncology patients.^27, 31^ Older patients are more likely to experience taste changes,^40^ reduced appetite,^41^ difficulty chewing and swallowing,^42^ and reduced functional capacity.^43^ Collectively, these factors can reduce the ability to meet nutritional needs. The association between age with MST, but not NUTRISCORE, may reflect that older individuals are more prone to weight loss and anorexia,^44, 45^ which are key components of MST scoring criteria. Conversely, NUTRISCORE also considers cancer type and treatment, which may be less directly tied to age.

Chemotherapy and radiation were also risk factors for malnutrition, as previously established.^2, 46, 47^ Side effects of these cancer treatments include nausea and vomiting, altered taste and smell, reduced appetite, gastrointestinal distress, dry mouth, and pain with oral intake, all of which can drive malnutrition.^2^ In contrast, hormone therapy had an inverse association with malnutrition risk that was statistically significant using the NUTRISCORE criteria. Subanalyses indicated that the cancer types most often receiving hormone therapy in the MACS cohort were endocrine cancer (primarily thyroid cancer) and breast cancer. The inverse association of hormone therapy with malnutrition risk could potentially be related to the overall low rates of malnutrition among these cancer types.

Obesity was inversely associated with risk for malnutrition measured by MST in our study. This finding aligns with prior research indicating BMI is associated with malnutrition risk.^16, 28^ Results linking a higher BMI to reduced risk for malnutrition may reflect that malnutrition frequently goes undiagnosed among patients with higher BMIs, as muscle wasting and other signs of malnutrition are more difficult to detect among these patients.^48^ Indeed, higher BMI was not significantly associated with malnutrition risk per NUTRISCORE, which also considers objective criteria such as cancer type and treatment. Overall, screening for and treating malnutrition is essential, regardless of BMI.^49, 50^

Male sex was positively associated with malnutrition risk using NUTRISCORE criteria. Although prior studies have not identified male sex as a risk factor for malnutrition, there is evidence that male oncology patients are at increased risk for cachexia.^51^ Malnutrition and cachexia share nearly identical clinical presentation (e.g., anorexia, weight loss, and muscle wasting).^52–55^ As a result, patients flagged as at risk for malnutrition may have underlying cachexia. Moreover, certain cancer types are more prone to develop cachexia. Since NUTRISCORE considers cancer type in its scoring criteria, it may identify more male patients with malnutrition risk than MST.

Head and neck, gastrointestinal, and lung cancers had the highest prevalence of malnutrition risk in HCI outpatient clinics, aligning with prior studies.^16, 27, 28^ Cancers of the head and neck and gastrointestinal tract and the associated treatments affecting these cancer sites result in symptoms that impact oral intake, namely changes in taste or smell, constipation, diarrhea, difficulty swallowing, fatigue, mouth sores, nausea, pain, and vomiting.^2, 56^ Some of the cancers included in these major cancer type groupings, such as pancreatic cancer, tend to be more aggressive and diagnosed at more advanced stages, increasing risk for malnutrition.^57^ Unknown or ill-defined cancer types that represent unspecified metastatic neoplasms also had high prevalence of malnutrition, likely due to their advanced cancer staging.^58^ Malnutrition screening may, therefore, be particularly important for these patient populations.

Strengths of our study include use and comparison of two validated malnutrition screening tools, MST and NUTRISCORE, both of which were one of six recommended malnutrition screening tools per a 2024 American Society for Parenteral and Enteral Nutrition (**ASPEN**) systematic review.^59^ Furthermore, MST is the recommended malnutrition screening tool per the Academy of Nutrition and Dietetics (**AND**).^11^ Our study is one of only a few studies conducted in the US^16^ to assess the prevalence of malnutrition risk and risk factors associated with malnutrition risk across diverse cancer types in an ambulatory setting.

This study’s limitations include its cross-sectional nature, which prevents any conclusions regarding causality. Due to limitations in abstracting data from the EMR, we were restricted to scoring for chemotherapy, radiation, or both when applying the NUTRISCORE criteria and were unable to account for other cancer treatment types included in the original scoring system.^19^ This may have led to an underestimation of malnutrition risk using this tool. Race is a potential predictor of malnutrition risk,^16^ but our sample had limited racial diversity to evaluate it as a predictor. MST screening at HCI outpatient clinics is recommended but not mandatory, and melanoma, genitourinary, breast, and gynecologic/breast cancer outpatient clinics had inconsistent implementation of MST screening over the final six-month period included in our analysis, which may affect estimates of malnutrition risk period prevalence for some cancer types.

## Conclusion

Our study provides valuable insights into the prevalence of malnutrition risk overall and across cancer types in the Mountain West region. We identified potential risk factors for malnutrition, including advanced cancer stage and smoking status. Implementing validated malnutrition screening tools, such as MST and NUTRISCORE, at outpatient oncology clinics is critical for the early identification and treatment of malnutrition. Further research assessing the prevalence of malnutrition using validated malnutrition assessment tools will advance the understanding of malnutrition risk factors and help to direct malnutrition screening, assessment, and interventional research.

## Data Availability

All data produced in the present study are available upon reasonable request to the authors

## List of Abbreviations Used

AND: Academy of Nutrition and Dietetics
ASPEN: American Society for Parenteral and Enteral Nutrition
BMI: Body Mass Index
EMR: Electronic Medical Record
GLIM: Global Leadership Initiative on Malnutrition
HCI: Huntsman Cancer Institute
HIPAA: Health Insurance Portability and Accountability Act
ICD-O: International Classification of Diseases for Oncology
MACS: Malnutrition Among Cancer Survivors
MST: Malnutrition Screening Tool
NCP: Nutrition Care Process
pg-SGA: Patient-Generated Subjective Global Assessment
RISR: Research Informatics Shared Resource
RUCA: Rural-Urban Commuting Area
US: United States
WHO: World Health Organization

## Acknowledgements

We acknowledge the Huntsman Cancer Institute patients for their contributions to this research. Research reported in this publication utilized the Research Informatics Shared Resource at Huntsman Cancer Institute at the University of Utah and was supported by the National Cancer Institute of the National Institutes of Health under Award Number P30CA042014. The content is solely the responsibility of the authors and does not necessarily represent the official views of the NIH.

## Ethics Approval and Consent to Participate

This study complies with the Declaration of Helsinki. The study was deemed exempt from full Institutional Review Board review and consent to participate was deemed unnecessary by the University of Utah Institutional Review Board under 45 CFR 46.104(d)(4). Reference: www.hhs.gov/ohrp/humansubjects/guidance/45cfr46.html

## Consent for Publication

Not Applicable.

## Availability of Data and Materials

The datasets analyzed during the current study are available from the corresponding author on reasonable request.

## Authorship Contributions

Conceptualization, M.P., R.H, M.H.; methodology, M.P., R.H., M.H.; validation, M.P., K.W., A.M.C., A.C., A.S; formal analysis R.H., M.H.; writing – original draft preparation, R.H., M.H.; writing – review and editing, M.P., K.W., A.M.C., A.C., A.S., J.E, T.K.V.; visualization, R.H., M.H.; supervision, M.P., K.W., A.M.C., A.C., A.S, T.K.V. All authors have read and agreed to the published version of the manuscript.

